# NCT Precision Oncology Thesaurus Drugs – a Curated Database for Drugs, Drug Classes, and Drug Targets in Precision Cancer Medicine

**DOI:** 10.1101/2022.09.11.22279783

**Authors:** Simon Kreutzfeldt, Alexander Knurr, Daniel Hübschmann, Peter Horak, Stefan Fröhling

## Abstract

Implementation of precision cancer medicine requires structured reporting of clinical actionable molecular alterations. The growing number of targeted anticancer drugs in clinical use and development necessitates a hierarchical ontology that focuses on the interactions between drugs and their targets and the impact of drug classes. Here we report the development of NCT POT (National Center for Tumor Diseases Drug Precision Oncology Thesaurus) Drugs, a manually curated cancer drug class ontology that integrates multiple data sources. This easily accessible resource is part of the NCT POT framework and can be downloaded at https://github.com/TMO-HD/nct-thesaurus with detailed documentation.

---

The vision of precision oncology entails a shift towards (semi-)automated, data-driven workflows that guide treatment based on an individual patient’s clinical and molecular profile and leverage a growing *profile-therapy-outcome* knowledge space derived from continuous surveillance of all cancer patients. The decision-making process in molecular tumor boards (MTBs), where all layers of information collected on a patient are integrated, currently requires a rapid review of clinically relevant biomedical literature and relies on manually curated knowledge bases. These resources, such as CIViC^1^ and OncoKB^2^, link genetic variants to cancer treatment sensitivity in the context of a defined cancer type (variant-drug-entity interaction) and are based on available clinical and preclinical evidence. In clinical decision-making, knowledge of drug class effects is highly relevant because it expands available evidence supporting a recommendation and offers additional options for implementing a therapeutic strategy. This creates a need for grouping drugs into classes based on the similarity of their chemical structures, mechanisms of action, and pharmacologic effects. Once a drug class effect has been established in clinical trials, treatment decisions more frequently consider additional aspects of cancer therapy, such as a drug’s toxicity, route of administration, cost, and impact on quality of life. In precision oncology, such drug class-based decisions can increase the number of drugs considered and, thus, a patient’s chances of receiving effective treatment.

A key prerequisite for the scalability of precision oncology workflows are semantic data standards and ontology-based knowledge bases, particularly for drugs, drug classes, and corresponding molecular targets. Comprehensive drug lists and a drug class taxonomy are necessary for accurately collecting clinical follow-up data, which are indispensable for generating new therapeutic insights. At multiple stages of such workflows, especially when searching for published evidence, approvals, or clinical trials, there is a need to translate between drug targets, drugs, and drug classes. For example, when searching for all published evidence of therapies combining immune checkpoint and PARP inhibitors, it is mandatory to have comprehensive lists of these agents to obtain complete hit lists, as the drug classes themselves often are not a findable search term. Similarly, when searching for all clinical trials investigating claudin-18 isoform 2-directed therapies, it is informative to know the currently available drugs (zolbetuximab [antibody], AMG 910 [bispecific T-cell engager], and several chimeric antigen receptor [CAR] T-cell therapies), as claudin-18 isoform 2 is not always listed in the trial description. Furthermore, approval and manufacturer information on in- and off-label treatments or contact information for compassionate use programs are also highly useful.

Currently, more than 200 cancer drugs are approved by the United States Food and Drug Administration (FDA); this is a highly heterogeneous group of compounds, with 15 to 20 being added each year. While many can be assigned to established therapeutic concepts such as chemotherapy, hormone therapy, immunotherapy, or targeted therapy, an increasing number of drugs entering clinical use evade these classifications (e.g., antibody-drug conjugates, radiopeptides, CAR-T-cell therapies, or vaccines). Another level of complexity arises from the low on-target specificity of individual compounds, such as small-molecule inhibitors. This makes classifications dependent on a wide range of pharmacologic effects and requires careful curation. Several drug ontologies, which may also be part of larger ontologies, are used in clinical practice. Many focus on chemical structure and prescription information (The Drug Ontology),^3^ approved drugs with limited target classification (e.g., Anatomical Therapeutic Chemical Classification,^4^ FDA Established Pharmacologic Class,^5^ and Systematized Nomenclature of Medicine Clinical Terms),^6^ or drug-target-only classification (Drug Target Ontology).^7^ None of these focuses on drugs in clinical development or drug target/biomarker information.

While the nomenclature and classification of genes, genetic variants, and tumor types have been intensively developed in recent years, standardized drug classes and ontologies for precision oncology based on drug targets are lacking. Considering the application scenarios outlined above, the most comprehensive specialized cancer ontology available, National Cancer Institute (NCI) Thesaurus,^8^ has excellent coverage of drugs, including nearly all relevant trial and preclinical drugs. However, its drug class taxonomy may not sufficiently reflect the clinical perspective and pharmacodynamic aspects. For example, drug targets are incompletely assigned, trade names are not tagged, and information on manufacturer and approval status is unavailable. The databases provided by HemOnc.org,^9^ another platform dedicated to providing information on cancer therapies, are more comprehensive regarding drug class assignment and cover approvals, trade names, and manufacturer information. However, many drugs are missing, particularly those in clinical trials, e.g., CAR T-cell products, vaccines, and preclinical compounds, and only sparse molecular target information is provided.^10^ Other resources such as Drugbank^11^ or the FDA-Approved Drugs database^12^ also have strengths in certain areas but fall short in others. Finally, the Jackson Laboratory Clinical Knowledgebase^13^ offers one of the most comprehensive drug knowledge bases, but it is incomplete and only commercially available.

Given the increasing number and clinical importance of targeted cancer treatments, we developed NCT POT (National Center for Tumor Diseases Precision Oncology Thesaurus) Drugs to address the rapidly growing need for a comprehensive cancer drug class ontology guiding clinical decision-making based on drug-target interactions and pharmacodynamic equivalence. Through this effort, we aimed to expand the scope of existing taxonomies and create an open-access, manually curated drug class database that continuously evolves based on first-in-class approvals and clinical guidelines and has interoperability with other resources to enhance and streamline precision cancer medicine workflows.

## Development of a Curated Drug Class Database

NCT POT Drugs provides comprehensive and detailed information on precision oncology drugs in clinical and preclinical use, i.e., their molecular targets, pharmacodynamic classes, manufacturers, and approval status, including references to approval criteria. To this end, it integrates information from public databases and is automatically updated on a regular basis as well as continuously manually curated to integrate different sources and supplement missing data (Fig 1A). NCT POT Drugs is embedded in the larger concept of the NCT POT framework, which provides a clinical application-oriented ontology of data domains relevant to precision oncology workflows. NCT POT is developed at NCT Heidelberg by a team of precision oncologists, bioinformaticians, and medical informatics specialists involved in the MASTER (Molecularly Aided Stratification for Tumor Eradication Research) program of the National Center for Tumor Diseases (NCT), the German Cancer Research Center (DKFZ), and the German Cancer Consortium (DKTK).^14^ NCT POT Drugs combines data from NCI Thesaurus,^8^ HemOnco.org,^15^ the FDA Approved Drug Database,^12^ the Human Genome Organisation Gene Nomenclature Committee (HGNC),^16^ CIViC,^1^ Ensembl databases,^17^ and OncoTree^18^ and integrates a self-maintained drug class ontology.

**FIG 1.**
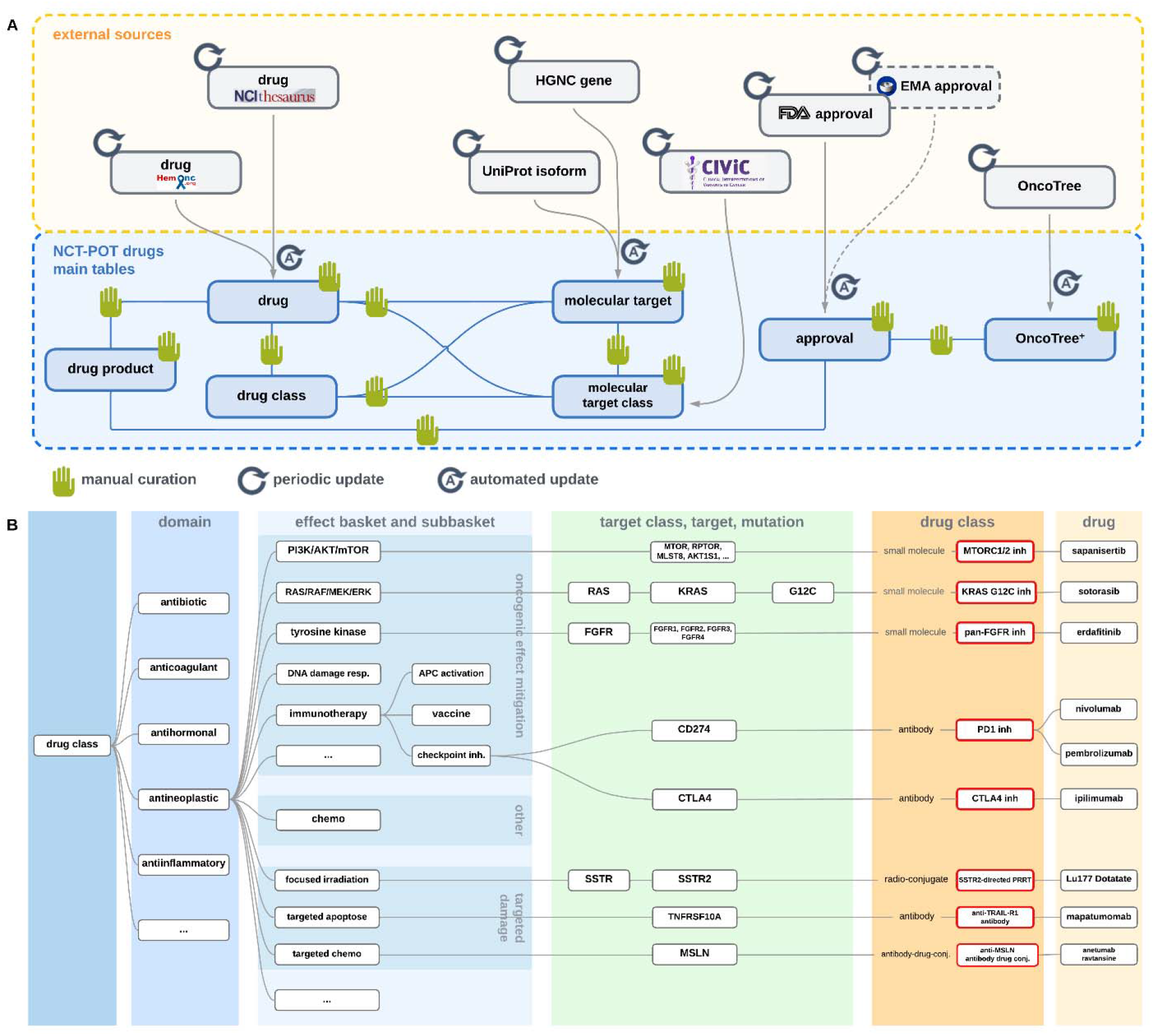
(A) Data sources integrated into NCT POT Drugs. Automated updates and manual curation steps increase data quality and keep the drug ontology up-to-date with changes in the underlying sources. (B) Drug class taxonomy. Domains separate drugs according to their main medical application. Effect baskets describe therapeutic strategies. Targets are sorted by protein families if appropriate, molecular targets, i.e., genes or gene products, and specific mutations. The resulting drug class is annotated with the molecule’s biochemical class and assigned to a specific drug. DDR, DNA damage response; APC, antigen-presenting cell; ICB, immune checkpoint blockade; inh, inhibitor.

Technically, NCT POT Drugs is implemented as a relational database consisting of the main entities *drug class, drug, molecular target class, molecular target, drug product, approval*, and *oncotree plus* (Fig 1A). There are mapping tables for n:m relations between *drug class* and *drug, drug class* and *molecular target*, and *drug* and *molecular target class*. The *drug class* entity has a hierarchical tree-like structure in which each drug class belongs to only one parent class. The *molecular target* category comprises genes indicated by their HGNC names; *molecular target class* includes information on drug-target interactions, gene variants such as resistance or sensitizing mutations, specific protein isoforms, or antibody binding epitopes; *drug product* covers commercial medicinal product information, including trade names, linked to the corresponding compounds; *drug approval* provides FDA and European Medicines Agency (EMA) approval information on drug entries in specific entities, as referenced in *oncotree plus*, which is a manually expanded version of OncoTree^18^ and also includes missing but clinically relevant diagnoses (such as triple-negative breast cancer).

Data integration and maintenance are conceptualized as a monthly, standard operating procedure-guided stepwise process (Fig 1A). It starts with downloads from the various sources and imports into source-specific tables, e.g., drugs from NCI Thesaurus, filtering for relevant data sets and building differentials to the already available imported tables. New entries or entries with updated attributes are then added or updated in the existing integrated table and marked “curation needed.” In the third step, all entries open for curation are reviewed, edited, and validated by a qualified curator who may add information, such as assignments to drug classes and drug targets. Besides these update cycles triggered by monthly resynchronization with the source databases, NCT POT Drugs is continuously edited when users encounter missing elements during their MTB case workup tasks, e.g., when searching for clinical trials or newly published evidence. The final curated dataset consists of seven main tables with six to 26 headers, comprising 18,262 drugs. Of these, 231 have been assigned to cancer-relevant drug classes in eight domains and 15 therapeutically relevant baskets (Fig 2B). From the 19,234 molecular targets, 64 have been assigned to specific target classes (Fig 2B).

NCT POT Drugs is implemented in the MTB of the NCT/DKFZ/DKTK MASTER program, providing information on drug class effects and pharmacologic equivalence between different drugs to support clinical decision-making. All data from NCT POT Drugs, including comprehensive documentation and download options, are available on Github (http://github.com/TMO-HD/NCT-POT/Drugs) under the XXX license. They are provided as an archive of tab-separated CSV files to facilitate technical implementation. Furthermore, we provide XML files for integration into the clinical documentation platform Onkostar®, which is widely used in Germany.

## Future Developments and Outlook

NCT POT Drugs is used in the MTB of the multicenter NCT/DKFZ/DKTK MASTER precision oncology program^14^ for young adults with advanced cancer and patients with rare malignancies. The data are used locally in MTB-supporting tools, such as the in-house-developed MTB curation, organization, and presentation platform Knowledge Connector (manuscript in preparation), where they support the search for published evidence or suitable clinical trials, while the drug class catalogs are provided, among others, to the clinical cancer registry (Onkostar®). Beyond local use, the implementation of NCT POT Drugs in precision cancer medicine networks across Germany is ongoing. Future plans to advance the concept of NCT POT Drugs include the integration of treatment regimens, improved annotation of molecular markers for drug approvals, the automated inclusion of new EMA approvals, and incorporation of pharmacogenomics data and preclinical information on drug-target interactions. Finally, we aim to provide the database in other technical formats such as XML, OBO, and OWL.

The rapidly growing amount of biological data from cancer patients and biomedical literature related to comprehensive molecular profiling of virtually all cancer entities has led to a situation referred to as “biomedical information overload.” Structuring data and creating formal ontologies can help with information processing and automation, reducing the burden of analysis for scientific and clinical purposes. An effective search, retrieval, analysis, and presentation of data in the context of MTBs requires automated data capture as well as manual curation of relevant ontology terms to facilitate interpretation and further processing. This novel precision oncology resource informs medical decision-making by providing therapy alternatives according to drug class effects based on biomarker information in the context of a given disease, allowing analysis of cross-entity and cross-biomarker efficacy of specific drugs and drug classes.

## Data Availability

All data produced are available online at http://github.com/TMO-HD/NCT-POT/Drugs

